# Association of pre-pandemic high-density lipoprotein cholesterol with risk of COVID-19 hospitalisation and death: the UK Biobank cohort study

**DOI:** 10.1101/2021.01.20.21250152

**Authors:** Camille Lassale, Mark Hamer, Álvaro Hernáez, Catharine R. Gale, G. David Batty

## Abstract

**Objective:** There is growing evidence of, and biological plausibility for, elevated levels of high-density lipoprotein cholesterol (HDL-C), being related to lower rates of severe infection. Accordingly, we tested whether pre-pandemic HDL-C within the normal range is associated with subsequent COVID-19 hospitalisations and death.

**Approach:** We analysed data on 317,306 participants from UK Biobank, a prospective cohort study, baseline data for which were collected between 2006 and 2010. Follow-up for COVID-19 was via hospitalisation records and a national mortality registry.

**Results:** After controlling for a series of confounding factors which included health behaviours, inflammatory markers, and socio-economic status, higher levels of HDL-C were related to a lower risk of later hospitalisation for COVID-19. The effect was linear (p-value for trend 0.001) such that a 0.2 mmol/L increase in HDL-C was associated with a corresponding 9% reduction in risk (odds ratio; 95% confidence interval: 0.91; 0.86, 0.96). A very similar pattern of association was apparent when COVID-19 mortality was the outcome of interest (odds ratio per 0.2 mmol/l increase in HDL-C: 0.90; 0.81, 1.00); again, a stepwise effect was evident (p-value for trend 0.03).

**Conclusions:** These novel results for HDL-C and COVID-19 events warrant testing in other studies.

## Introduction

High-density lipoprotein cholesterol (HDL-C) has traditionally been linked with coronary heart disease and stroke.^1^ While conventional epidemiological studies consistently demonstrate that elevated levels of this cholesterol fraction confer protection against vascular events,^2^ support for such a gradient has been lacking in people genetically predisposed to low concentrations of HDL-C,^3^ Mendelian randomisation studies,^4^ and randomized clinical trials utilising HDL-C–elevating medication.^5^

More recently, HDL-C has been implicated in the pathogenesis of other health endpoints, including infectious disease. While most investigators testing the link between HDL-C and infection have done so in prognostic studies of patients groups,^8^ in one of the few cohort analyses of apparently healthy individuals, people with lower levels of baseline HDL-C experienced a greater subsequent risk of hospitalisation for gastroenteritis, urinary tract infection, and bacterial pneumonia.^9^ Plausible mechanisms include the HDL-C–mediated sequestration of pathogen-associated lipids, and regulation of immune cells proliferation, maturation, and function which results in neutralization or clearance of pro-inflammatory endotoxins.^7^

This evidence base raises the possibility of a link between HDL-C and COVID-19, the disease caused by severe acute respiratory syndrome coronavirus 2. Using UK Biobank, a prospective cohort study, we have recently shown that an unfavourable pre-pandemic vascular risk factor profile – low HDL-C included – is associated with a higher risk of hospitalization for COVID-19.^10^ Whether HDL-C across the normal range offers predictive capacity for COVID-19 hospitalisations is, however, untested, and this is the purpose of the present study. Further, as the present pandemic has unfolded, this cohort has accumulated sufficient deaths from this disease to facilitate analyses with the aim of corroborating any associations with hospitalisations.

## Methods

### Study population

We used data from the UK Biobank, a prospective cohort study,^11^ baseline data collection for which took place between 2006 and 2010 across centres in the UK, yielding a sample of 502,655 people (448,919 from England) aged 40-69 years. Ethical approval was provided by the North-West Multi-centre Research Ethics Committee (11/NW/0382; 16/NW/0274).

### Baseline data collection

At baseline, non-fasting venous blood samples were drawn and assayed for total cholesterol, HDL-C, and triglycerides using a Beckman Coulter AU5800 analytical platform. Low density lipoprotein (LDL)-cholesterol values were calculated using the Friedewald equation.^12^ Total blood count (leukocyte, platelet, haemoglobin) as markers of immune function were analysed using an automated Coulter LH 750. Physician-diagnosed cardiovascular disease (heart attack, angina, stroke), diabetes, cholesterol-lowering drugs use, cigarette smoking, alcohol intake, highest educational attainment, ethnicity,^13^ number of people living in the household, and physical activity in the prior month were self-reported using standard enquiries.^14^ Body mass index was computed using direct measurements of height and weight using the usual formulae.^15^ Hypertension was defined as elevated measured blood pressure (≥140/90 mmHg) and/or use of anti-hypertensive medication. Townsend index of neighbourhood deprivation was based on postcode linkage.^14^ Provided by Public Health England, data on COVID-19 status in hospitalised patients in England covered the period 16^th^ March until 31^st^ May 2020. Tests were performed in accredited laboratories on samples from combined nose/throat swabs using real time polymerase chain reaction. Participants were also linked to long-standing national mortality records from which death from COVID-19, our outcome of interest, for the period 1^st^ March to 30^th^ September 2020, was denoted by the emergency International Classification of Disease (version 10) code U07.1 (COVID-19, virus identified).

### Statistical analyses

We used logistic regression analyses to compute odds ratios with accompanying 95% confidence intervals to summarise the relation between HDL-C and later COVID-19 hospitalisation or death. In a first analytical approach, we created an HDL-C variable with eight categories which was designed to examine the shape of the HDL-C–COVID-19 relationship (<1.0 [referent], 1.0 to <1.2, 1.2 to <1.4, 1.4 to <1.6, 1.6 to <1.8, 1.8 to <2.0, 2.0 to <2.2, ≥2.2 mmol/L); in the analyses of COVID-19 deaths, the latter two categories were collapsed owing to a lower number of events. With preliminary analyses suggesting a linear gradient, we were then able to summarise the relationship for a unit change in HDL-C (0.2 mmol/L increase). In taking both approaches, we first adjusted for age and sex (comparator model) and then, in the multivariable model, a series of covariates which included inflammatory markers, lifestyle factors, and socioeconomic circumstances.

## Results

In 317,306 (171,466 women) participants with complete data on baseline covariates, there were 869 hospitalisations for COVID-19 (427 in women) during the surveillance period. As illustrated in figure 1, in age- and sex-adjusted analyses, relative to the group with the lowest concentration of HDL-C, those in the highest experienced around one third of the risk of hospitalization for COVID-19 (odds ratio; 95% confidence interval: 0.31; 0.19, 0.52). There was also evidence of a stepwise relationship (p-value linear trend <0.001) such that lower disease risk was apparent in people with higher level of this cholesterol fraction. Summarising this trend, a 0.2 mmol/L increase in HDL-C was associated with a 15% lower risk of subsequent hospitalisation (0.85; 0.82, 0.89). Adjusting for an array of confounding factors – health behaviours, inflammatory markers, and socio-economic status – resulted in a shallower HDL-C–COVID-19 gradient but the linear association remained (p for linear trend 0.001) with 0.2 mmol/L increase in the cholesterol fraction now corresponding to a 9% lower risk of the disease (0.91; 0.86, 0.96).

**Figure 1.**
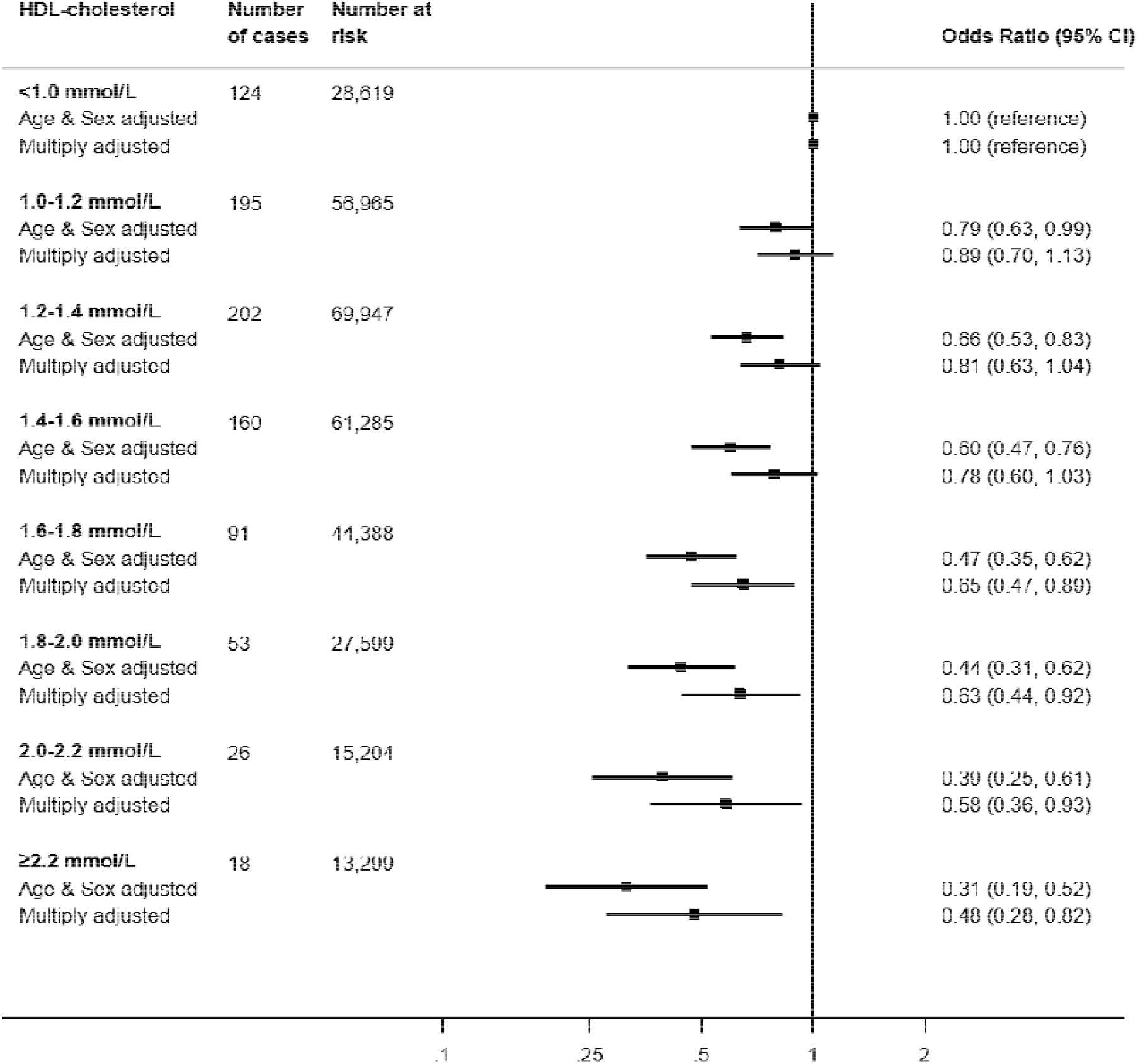
HDL-C concentration (2006-10) and risk of subsequent hospitalisation for COVID-19 (2020) in UK Biobank (N=317,306) Multiply adjusted odds ratios are adjusted for age, sex, ethnicity, education, number in household, area deprivation, body mass index, leisure time physical activity, alcohol intake, smoking habit, diagnosed diabetes, cardiovascular disease, or hypertension, cholesterol-lowering medication, LDL-cholesterol, triglycerides, haemoglobin, white blood cell, and platelet count.

Next, we tested the results for HDL-C and COVID-19 mortality in 317,827 participants (171,675 women) in whom there were 232 COVID-19 deaths (79 in women) during the surveillance period. As apparent in figure 2, the HDL-C relation with mortality was similar to that seen in the prior analyses for hospitalisations. Thus, after controlling for age and sex, relative to the group with the lowest concentration of HDL-C, those individuals in the group with the highest HDL-C concentration (≥2.0 mmol/L) had less than half of the risk of death ascribed to COVID-19 (odds ratio; 95% confidence interval: 0.40; 0.21, 0.76). Again, there was evidence of a dose-response effect across the full range of HDL-C values (p-value for trend 0.0003) whereby an increase in HDL-C of 0.2 mmol/L produced a 15% lower risk of mortality (0.85; 0.78, 0.92). As apparent from the breadth of the confidence intervals for several point estimates, however, some of these analyses had lower precision owing to the lower number of deaths. Although multiple adjustment for covariates led to attenuation of the magnitude of the HDL-C–death relation, the shape apparent in the minimally-adjusted analyses was retained (p-value for trend 0.03); an increase of 0.2 mmol/L was associated with a 10% lower risk (0.90; 0.81, 1.00).

**Figure 2.**
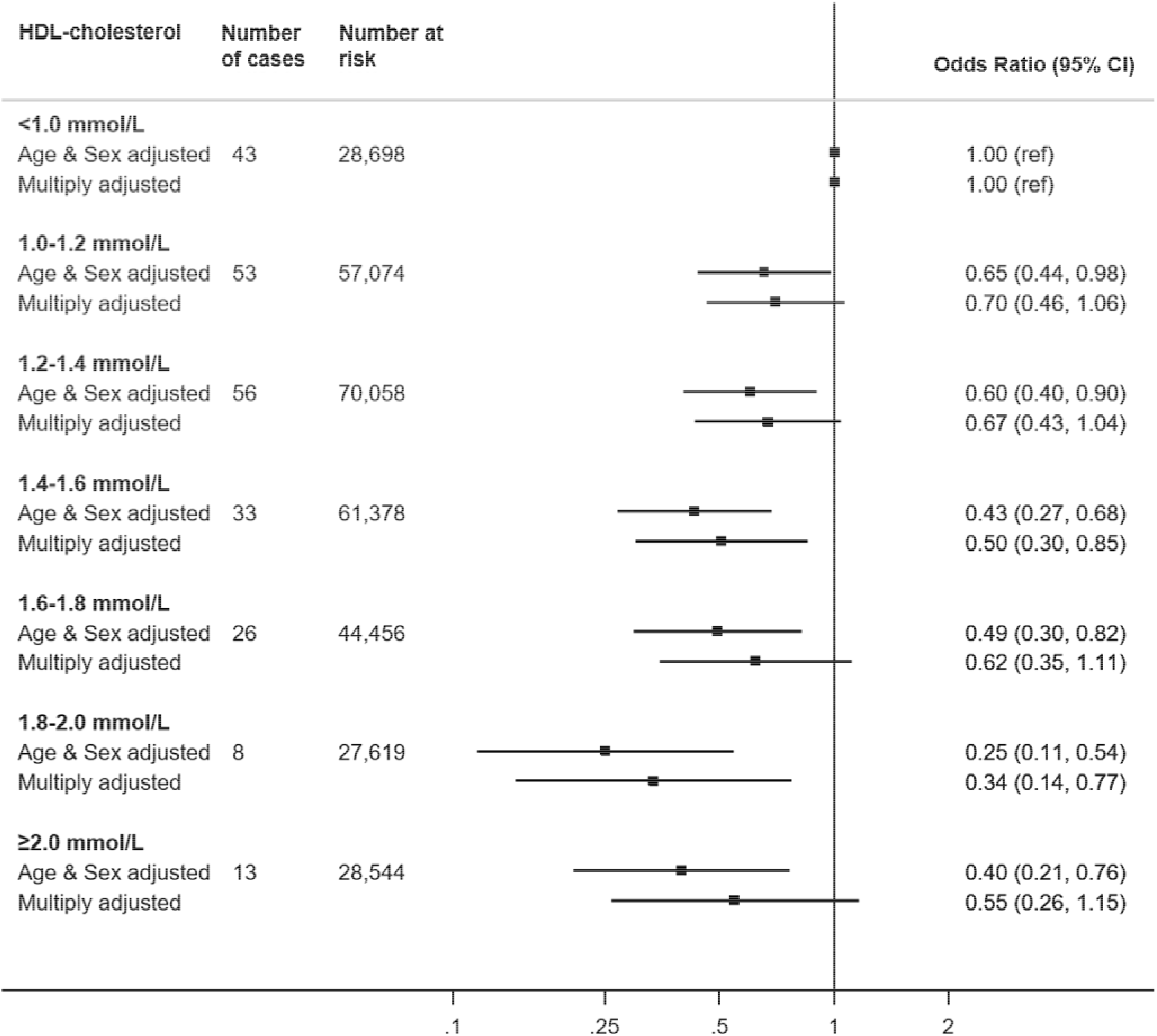
HDL-C concentration (2006-10) and risk of death from COVID-19 (2020) in UK Biobank (N=317,827) Multiply adjusted odds ratios are adjusted for age, sex, ethnicity, education number in household, area deprivation, body mass index, leisure time physical activity, alcohol intake, smoking habit, diagnosed diabetes, cardiovascular disease, or hypertension, cholesterol-lowering medication, LDL-cholesterol, triglycerides, haemoglobin, white blood cell, and platelet count.

## Discussion

To the best of our knowledge, this is the first study to examine the shape of the relationship between pre-pandemic HDL-C levels and risk of later COVID-19 events. Our salient finding was that, net of an array of confounding factors, higher concentrations of HDL-C were associated with protection against hospitalisation for, and death from, the disease. That COVID-19 events ascertained using different approaches revealed very similar effects increasing confidence in our novel results. In other analyses from the present dataset we have reported what are now regarded as established associations of classic vascular risk factors such as raised levels of glycosylated haemoglobin,^16^ body weight,^15^ and blood pressure^10^ with COVID-19, as apparent in studies based in the US,^17^ Italy,^18^ China,^19, 20^ and Brazil.^21^ We have also shown higher rates of hospitalisation for the disease in people of increased age, male sex, socioeconomic disadvantage,^14^ and ethnic minority groups.^13^

### Comparison with existing studies

Levels of HDL-C appear to change in the presence of coronavirus disease 2019 (COVID-19), such that, relative to healthy controls, patients with COVID-19 have lower levels of HDL-C concentrations in conjunction with acute elevation in systemic inflammatory markers.^22, 23^ While a lowering of HDL-C levels appears to be a consequence of COVID-19, the reverse, whereby pre-pandemic HDL-C levels may offer some predictive capacity for this disease, has been little examined. Taking a Mendelian Randomisation approach, investigators using UK Biobank data have shown an inverse association between baseline HDL-C and hospitalisations for any infectious disease up to 10 years later.^24^ In contrast, using a conventional epidemiological study design, analyses based on two Danish community-based, prospective cohort studies with up to 20 years of follow-up revealed a ‘U’-shaped relationship whereby the greatest risk of hospitalisation for any infection was apparent at opposing ends of the HDL-C continuum.^9^ There was no clear evidence of such a quadratic effect in the present study. Studies linking with respiratory disease and earlier measurement of HDL-C are scarce and we are unaware of any other analyses with COVID-19 as the endpoint.

### Mechanisms of effect

Potential mechanisms for the HDL-C–COVID-19 gradient include the anti-inflammatory properties of the lipoprotein which are related to lower risk of cardiovascular disease in humans,^25^ particularly relevant in COVID-19 complications. Further, HDL-C can act as a direct modulator of immunity by inhibiting haematopoietic stem cells proliferation and affecting the activity and function of immune cells by changing the cholesterol content of the lipid rafts in immune cell membranes.^7^ A direct binding and neutralization of viral particles by HDL-C may also underpin the observed association.^26^

### Study strengths and weaknesses

The strengths of this study include the measurement of biomarkers that preceded the onset of COVID-19, so ruling out reverse causality. While large, the study sample is also well-characterised. That UK Biobank participants represent only 6% of the target population, however, means that the present data cannot be used to estimate prevalence or incidence in the general population, although established risk factor associations appear generalisable.^27^ HDL-C levels were measured up to 14 years before COVID-19 case assessment and this raises concerns about their utility for current values, however, in a reassessment a mean of 4.4 years after baseline examination they showed high test-retest stability (correlation coefficient 0.85, p<0.001) in a subsample (N=13,430). While the HDL-C–COVID-19 gradient was robust to the adjustment of various covariates, it is plausible that unmeasured confounding factors might explain the association. To this extent, because the data are observational, we cannot be dogmatic about causality. Further scrutiny of our results, including the application of the Mendelian Randomisation approach where a genetic proxy for HDL-C is used as the exposure of interest, is required.

In conclusion, the novel association between higher levels of HDL-C and lower risk of hospitalisation and death due to COVID-19 warrant testing in using other study designs.

## Data Availability

Data sharing: Data from UK Biobank (https://www.ukbiobank.ac.uk/) are available to bona fide researchers on application. Part of this research has been conducted using the UK Biobank Resource under Application 10279.

## References

1. Gordon DJ, Probstfield JL, Garrison RJ, Neaton JD, Castelli WP, Knoke JD, Jacobs DR, Jr., Bangdiwala S, Tyroler HA. High-density lipoprotein cholesterol and cardiovascular disease. Four prospective american studies. Circulation. 1989;79:8–15

2. Di Angelantonio E, Sarwar N, Perry P, Kaptoge S, Ray KK, Thompson A, Wood AM, Lewington S, Sattar N, Packard CJ, Collins R, Thompson SG, Danesh J. Major lipids, apolipoproteins, and risk of vascular disease. Jama. 2009;302:1993–2000

3. Frikke-Schmidt R, Nordestgaard BG, Stene MC, Sethi AA, Remaley AT, Schnohr P, Grande P, Tybjaerg-Hansen A. Association of loss-of-function mutations in the abca1 gene with high-density lipoprotein cholesterol levels and risk of ischemic heart disease. Jama. 2008;299:2524–2532

4. Holmes MV, Asselbergs FW, Palmer TM, Drenos F, Lanktree MB, Nelson CP, Dale CE, Padmanabhan S, Finan C, Swerdlow DI, Tragante V, van Iperen EP, Sivapalaratnam S, Shah S, Elbers CC, Shah T, Engmann J, Giambartolomei C, White J, Zabaneh D, Sofat R, McLachlan S, Doevendans PA, Balmforth AJ, Hall AS, North KE, Almoguera B, Hoogeveen RC, Cushman M, Fornage M, Patel SR, Redline S, Siscovick DS, Tsai MY, Karczewski KJ, Hofker MH, Verschuren WM, Bots ML, van der Schouw YT, Melander O, Dominiczak AF, Morris R, Ben-Shlomo Y, Price J, Kumari M, Baumert J, Peters A, Thorand B, Koenig W, Gaunt TR, Humphries SE, Clarke R, Watkins H, Farrall M, Wilson JG, Rich SS, de Bakker PI, Lange LA, Davey Smith G, Reiner AP, Talmud PJ, Kivimäki M, Lawlor DA, Dudbridge F, Samani NJ, Keating BJ, Hingorani AD, Casas JP. Mendelian randomization of blood lipids for coronary heart disease. Eur Heart J. 2015;36:539–550

5. Keene D, Price C, Shun-Shin MJ, Francis DP. Effect on cardiovascular risk of high density lipoprotein targeted drug treatments niacin, fibrates, and cetp inhibitors: Meta-analysis of randomised controlled trials including 117,411 patients. Bmj. 2014;349:g4379

6. Hamer M, Batty GD, Kivimaki M. Obesity, metabolic health, and history of cytomegalovirus infection in the general population. J Clin Endocrinol Metab. 2016;101:1680–1685

7. Catapano AL, Pirillo A, Bonacina F, Norata GD. Hdl in innate and adaptive immunity. Cardiovasc Res. 2014;103:372–383

8. Shor R, Wainstein J, Oz D, Boaz M, Matas Z, Fux A, Halabe A. Low hdl levels and the risk of death, sepsis and malignancy. Clin Res Cardiol. 2008;97:227–233

9. Madsen CM, Varbo A, Tybjærg-Hansen A, Frikke-Schmidt R, Nordestgaard BG. Ushaped relationship of hdl and risk of infectious disease: Two prospective population-based cohort studies. European heart journal. 2018;39:1181–1190

10. Batty GD, Hamer M. Vascular risk factors, framingham risk score, and covid-19: Community-based cohort study. Cardiovasc Res. 2020;116:1664–1665

11. Sudlow C, Gallacher J, Allen N, Beral V, Burton P, Danesh J, Downey P, Elliott P, Green J, Landray M, Liu B, Matthews P, Ong G, Pell J, Silman A, Young A, Sprosen T, Peakman T, Collins R. Uk biobank: An open access resource for identifying the causes of a wide range of complex diseases of middle and old age. PLoS Med. 2015;12:e1001779

12. Friedewald WT, Levy RI, Fredrickson DS. Estimation of the concentration of low-density lipoprotein cholesterol in plasma, without use of the preparative ultracentrifuge. Clinical chemistry. 1972;18:499–502

13. Lassale C, Gaye B, Hamer M, Gale CR, Batty GD. Ethnic disparities in hospitalisation for covid-19 in england: The role of socioeconomic factors, mental health, and inflammatory and pro-inflammatory factors in a community-based cohort study. Brain Behav Immun. 2020;88:44–49

14. Batty GD, Deary IJ, Luciano M, Altschul DM, Kivimaki M, Gale CR. Psychosocial factors and hospitalisations for covid-19: Prospective cohort study based on a community sample. Brain Behav Immun. 2020;89:569–578

15. Hamer M, Gale CR, Kivimaki M, Batty GD. Overweight, obesity, and risk of hospitalization for covid-19: A community-based cohort study of adults in the united kingdom. Proc Natl Acad Sci U S A. 2020;117:21011–21013

16. Hamer M, Gale CR, Batty GD. Diabetes, glycaemic control, and risk of covid-19 hospitalisation: Population-based, prospective cohort study. Metabolism. 2020;112:154344

17. Richardson S, Hirsch JS, Narasimhan M, Crawford JM, McGinn T, Davidson KW, Barnaby DP, Becker LB, Chelico JD, Cohen SL, Cookingham J, Coppa K, Diefenbach MA, Dominello AJ, Duer-Hefele J, Falzon L, Gitlin J, Hajizadeh N, Harvin TG, Hirschwerk DA, Kim EJ, Kozel ZM, Marrast LM, Mogavero JN, Osorio GA, Qiu M, Zanos TP. Presenting characteristics, comorbidities, and outcomes among 5700 patients hospitalized with covid-19 in the new york city area. Jama. 2020

18. Grasselli G, Greco M, Zanella A, Albano G, Antonelli M, Bellani G, Bonanomi E, Cabrini L, Carlesso E, Castelli G, Cattaneo S, Cereda D, Colombo S, Coluccello A, Crescini G, Forastieri Molinari A, Foti G, Fumagalli R, Iotti GA, Langer T, Latronico N, Lorini FL, Mojoli F, Natalini G, Pessina CM, Ranieri VM, Rech R, Scudeller L, Rosano A, Storti E, Thompson BT, Tirani M, Villani PG, Pesenti A, Cecconi M. Risk factors associated with mortality among patients with covid-19 in intensive care units in lombardy, italy. JAMA Intern Med. 2020;180:1345–1355

19. Wu C, Chen X, Cai Y, Xia J, Zhou X, Xu S, Huang H, Zhang L, Zhou X, Du C, Zhang Y, Song J, Wang S, Chao Y, Yang Z, Xu J, Zhou X, Chen D, Xiong W, Xu L, Zhou F, Jiang J, Bai C, Zheng J, Song Y. Risk factors associated with acute respiratory distress syndrome and death in patients with coronavirus disease 2019 pneumonia in wuhan, china. JAMA internal medicine. 2020

20. Zhou F, Yu T, Du R, Fan G, Liu Y, Liu Z, Xiang J, Wang Y, Song B, Gu X, Guan L, Wei Y, Li H, Wu X, Xu J, Tu S, Zhang Y, Chen H, Cao B. Clinical course and risk factors for mortality of adult inpatients with covid-19 in wuhan, china: A retrospective cohort study. Lancet. 2020;395:1054–1062

21. Baqui P, Bica I, Marra V, Ercole A, van der Schaar M. Ethnic and regional variations in hospital mortality from covid-19 in brazil: A cross-sectional observational study. Lancet Glob Health. 2020;8:e1018–e1026

22. Wei X, Zeng W, Su J, Wan H, Yu X, Cao X, Tan W, Wang H. Hypolipidemia is associated with the severity of covid-19. Journal of clinical lipidology. 2020;14:297–304

23. Sorokin AV, Karathanasis SK, Yang ZH, Freeman L, Kotani K, Remaley AT. Covid-19—associated dyslipidemia: Implications for mechanism of impaired resolution and novel therapeutic approaches. The FASEB Journal. 2020;34:9843–9853

24. Trinder M, Walley KR, Boyd JH, Brunham LR. Causal inference for genetically determined levels of high-density lipoprotein cholesterol and risk of infectious disease. Arteriosclerosis, thrombosis, and vascular biology. 2020;40:267–278

25. Soria-Florido MT, Schröder H, Grau M, Fitó M, Lassale C. High density lipoprotein functionality and cardiovascular events and mortality: A systematic review and meta-analysis. Atherosclerosis. 2020;302:36–42

26. Kane JP, Hardman DA, Dimpfl JC, Levy JA. Apolipoprotein is responsible for neutralization of xenotropic type c virus by mouse serum. Proceedings of the National Academy of Sciences. 1979;76:5957–5961

27. Batty GD, Gale CR, Kivimaki M, Deary IJ, Bell S. Comparison of risk factor associations in uk biobank against representative, general population based studies with conventional response rates: Prospective cohort study and individual participant meta-analysis. BMJ. 2020;368:m131

